# OVERVIEW OF RENAL OSTEODYSTROPHY IN BRAZIL: A CROSS-SECTIONAL STUDY

**DOI:** 10.1101/2022.09.29.22280450

**Authors:** Cinthia E. M. Carbonara, Noemi A. V. Roza, Luciene M. dos Reis, Aluízio B. Carvalho, Vanda Jorgetti, Rodrigo Bueno de Oliveira

## Abstract

**Introduction:** Epidemiologic profile of renal osteodystrophy (ROD) is changing over time and cross-sectional studies provide essential information to improve care and health policies. The *Brazilian Registry of Bone Biopsy* (REBRABO) is a prospective, national-multicenter cohort that aims to provide clinical information on ROD.

**Methods:** From Aug/2015-Dec/2021, clinical-demographic, laboratory and ROD information were included.

**Results:** Data from 386 individuals were considered. Age was 52 (42–60) years; 198 (51%) were male; 315 (82%) were on hemodialysis; osteitis fibrosa (OF) [163 (42%)] and adynamic bone disease (ABD) [96 (25%)] were the most prevalent forms of ROD; 190 (49%) had the diagnosis of osteoporosis, 82 (56%) vascular calcification; 138 (36%) bone aluminum accumulation and 137 (36%) iron intoxication; patients with high turnover were prone to present a higher prevalence of symptoms.

**Conclusions:** Elevated proportion of patients were diagnosed with OF and ABD, as well as osteoporosis, vascular calcification and clinical symptoms.

## INTRODUCTION

Renal osteodystrophy (ROD) is a common complication of chronic kidney disease (CKD) associated with bone fractures, vascular calcification, and decreased quality of life.^1-3^ In the last 40 years, the availability of new drugs and improvements in dialysis treatments have changed the epidemiologic profile of ROD.^4-6^ Some authors observed a dominant prevalence of adynamic bone disease (ABD), while others reported a predominance of osteitis fibrosa (OF).^7,8^

The reports of case series and cohorts involving patients with ROD depict geography and ethnic differences,^7,8^ which may also be related to disparities in treatment access and heterogeneous standards of quality in provided care.^9-11^ Regional information related to ROD may be important to support changes in health policies and to recognize important local patterns.

The *Brazilian Registry of Bone Biopsy* (REBRABO) is a prospective, national multicenter cohort that aims to provide clinical information on ROD.^12^This brief communication represents an update from previously published data from REBRABO.^8^ The main objective of this subanalysis was to describe the profile of ROD, including clinically relevant associations. The secondary objective was to explore regional differences in ROD.

## MATERIALS AND METHODS

This study was conducted as a subanalysis of REBRABO. During the period from Aug 2015 to Dec 2021, 511 patients were included in REBRABO. Exclusion criteria were: no bone biopsy report (N=40), GFR > 90 mL/min (N=28), not signed consent (N=24), bone fragments inadequate for diagnosis (N=23), bone biopsy indicated by a specialty other than nephrology (N=6), and < 18 years old (N=4). The local ethics committee approved the study protocol (CAAE 4131141.6.0000.5404), and the research activities being reported are consistent with the Declaration of Helsinki.

All clinical, demographic and laboratory data were collected in reference to bone biopsy date using standard electronic forms available at the REBRABO web system. The baseline data were entered by a nephrologist who performed the bone biopsy and were validated by a single researcher. The following data were considered: age, sex, ethnicity, CKD etiology, dialysis vintage and modality, comorbidities, symptoms and complications related to ROD, drugs related to CKD-MBD, serum levels of total calcium, phosphate, parathormone, alkaline phosphatase, 25-hydroxyvitamin D, and hemoglobin. We considered the recommended range for serum levels as follows: calcium (8.8-10.2 mg/dL), phosphate (3.5-5.0 mg/dL), parathormone (130-565 pg/mL) and 25-vitD (30-60 ng/mL).

Bone fragments were obtained via transiliac bone biopsies using an electrical trephine after prelabeling with tetracycline (3 days) administered over two separate periods. Undecalcified bone fragments were submitted to standard processing for histological studies.^13^ Bone sections were stained with toluidine blue. Al bone content was identified by solochrome azurine staining, and iron was identified by Pearls staining. Al accumulation or iron intoxication was considered when ≥30% of the surface was covered. The samples from individual patients were classified as having OF, mixed uremic osteodystrophy (MUO), ABD, osteomalacia (OM), normal/minor alterations, osteoporosis, bone aluminum (Al) accumulation and iron intoxication.

Continuous variables are reported as the means ± SDs or medians and interquartile intervals. Categorical data are reported as frequencies and percentages. The Mann-Whitney test and X^2^ test were applied for comparisons. Statistical analyses were performed using SPSS 22.0 (SPSS Inc., Chicago, IL). A two-sided p value < 0.05 was considered statistically significant.

## RESULTS

Data from 386 individuals were considered in our analysis. The mean age was 52 (42–60) years; 198 (51%) were male and 160 (41%) Caucasian; 315 (82%) were on hemodialysis, 31 (8%) on peritoneal dialysis, and 40 (10%) on conservative management; 73 (19%) patients had undergone parathyroidectomy; 236 (61%) of patients were in use of sevelamer hydrochloride, 104 (27%) calcium salts, 104 (27%) vitamin D receptor activators, 66 (17%) native vitamin D, and 98 (25%) cinacalcet hydrochloride (Table 1).

**Table 1.** General clinical and biochemical data.

|  | N = 386 |
| --- | --- |
| Age (years) | 52 (42–60) |
| Body mass index (kg/m<sup>2</sup>) | 24.1 (22–27) |
| Gender (male; N, %) | 198 (51) |
| Ethnicity (Caucasian; N, %) | 160 (41) |
| Diabetes mellitus (N, %) | 57 (15) |
| Prior cardiovascular disease (N, %) | 36 (9) |
| CKD etiology (N, %) |  |
| Hypertension | 105 (27) |
| Chronic glomerulonephritis | 94 (24) |
| Diabetes *mellitus* | 46 (12) |
| Dialysis vintage (months) | 84 (36–156) |
| Hemoglobin (g/dL) | 11.5 (10–13) |
| Total calcium (mg/dL) | 9.3 (8.6–9.9) |
| Phosphate (mg/dL) | 4.9 (3.9–6.2) |
| Parathormone (pg/mL) | 233 (63–783) |
| Alkaline phosphatase (IU/L) | 124 (79–225) |
| 25-vitamin D (ng/mL) | 29 (21–38) |

### ROD diagnosis

The main indications for bone biopsy were suspicion of bone Al accumulation in 133 (34%) patients, research protocol in 114 (29%), refractory bone pain in 55 (14%) and refractory hypercalcemia/phosphatemia in 33 (8%). OF [163 (42%) patients] and ABD [96 (25%) patients] were the most prevalent forms of ROD; 83 (21%) had MUO, 19 (5%) OM and 25 (6%) normal/minor alterations; 196 (52%) of patients had abnormal bone mineralization; 190 (49%) of patients had the diagnosis of osteoporosis; 31 (27%) of patients with low turnover bone disease had undergone parathyroidectomy; 138 (36%) of patients presented the diagnosis of bone Al accumulation and 137 (36%) iron intoxication (Figure 1).

**Figure 1.**
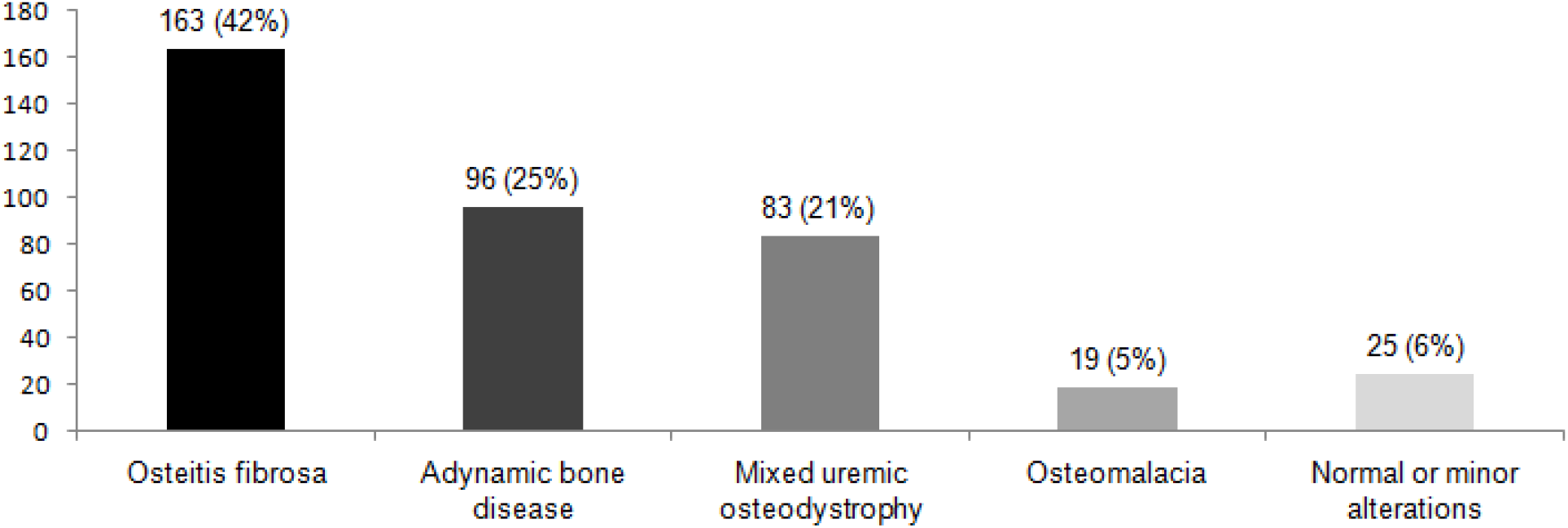
Prevalence of renal osteodystrophy.

### Prevalence of symptoms, vascular calcification and bone complications

A high prevalence of symptoms, vascular calcification and bone complications were detected. Patients with high turnover bone disease were prone to present a higher prevalence of weakness, bone pain, myalgia and itching than those with low turnover (Table 2). No differences in the prevalence of symptoms, vascular calcification and bone complications were noted according to bone volume or mineralization {exception for myalgia: patients with abnormal mineralization were prone to present more myalgia [69 (60%) *vs*. 46 (40%); p = 0.04]}.

**Table 2.**
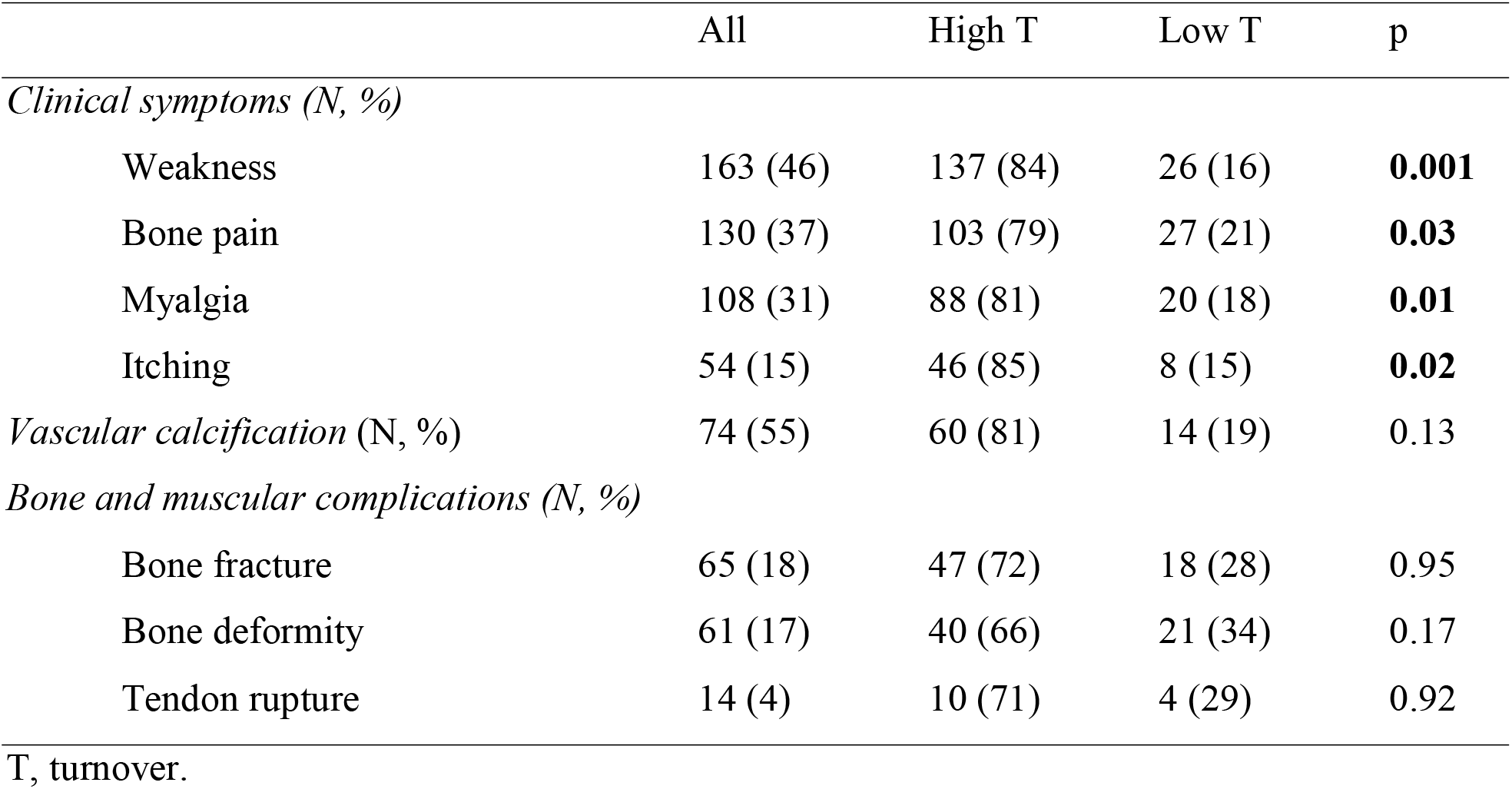
Prevalence of symptoms, vascular calcification and bone complications according to bone turnover.

|  | All | High T | Low T | p |
| --- | --- | --- | --- | --- |
| *Clinical symptoms (N, %)* |  |  |  |  |
| Weakness | 163 (46) | 137 (84) | 26 (16) | **0.001** |
| Bone pain | 130 (37) | 103 (79) | 27 (21) | **0.03** |
| Myalgia | 108 (31) | 88 (81) | 20 (18) | **0.01** |
| Itching | 54 (15) | 46 (85) | 8 (15) | **0.02** |
| *Vascular calcification (N, %)* | 74 (55) | 60 (81) | 14 (19) | 0.13 |
| *Bone and muscular complications (N, %)* |  |  |  |  |
| Bone fracture | 65 (18) | 47 (72) | 18 (28) | 0.95 |
| Bone deformity | 61 (17) | 40 (66) | 21 (34) | 0.17 |
| Tendon rupture | 14 (4) | 10 (71) | 4 (29) | 0.92 |

T, turnover.

### Effects of ROD on serum biomarkers

The proportion of patients who presented within the recommended range for serum levels of calcium was 210 (54%), phosphate 132 (34%), parathormone 116 (30%), and 25-hydroxy vitamin D 76 (43%). Hyperphosphatemia was observed in 185 (48%) patients, while hypercalcemia was observed in 60 (16%).

Patients with high turnover bone disease were prone to present a higher prevalence of serum P levels out of the recommended range compared to those with low turnover [186 (75%) *vs*. 61 (25%); p = 0.001]. Patients with abnormal bone mineralization were prone to present a lower prevalence of serum P levels out of the recommended range than those with normal mineralization [118 (47%) *vs*. 132 (53%); p = 0.007]. No other differences were noted according to bone turnover, mineralization and volume.

### The influence of geographic region on ROD

A total of 300 (78%) bone biopsies came from the Southeast Region, 74 (19%) from the Northeast, 8 (2%) from the North, 3 (1%) from the Midwest, and 1 (0.3%) from the South. The type of ROD, the prevalence of osteoporosis and iron intoxication did not change according to geographic region (p = 0.08, 0.45, and 0.36, respectively).

However, we observed a distinct occurrence of bone aluminum accumulation across the regions. All bone biopsies samples from North 8 (100%) presented this diagnosis, while the prevalence from Northeast bone biopsies was 31 (42%), Southeast 98 (33%) and Midwest 1 (33%) (p = 0.02).

## DISCUSSION

Our study shows the following findings: (1) OF and ABD were the most prevalent forms of ROD; (2) osteoporosis and vascular calcification were detected in almost half of the sample, while Al and iron deposition in bone reached more than one-third of patients; (3) patients with ROD, especially those with high turnover bone disease, experienced a high prevalence of clinical symptoms; and (4) bone Al accumulation occurred in a high proportion in patients from the North and Northeast Regions.

Compared with a previous report,^8^ there was a decrease in the prevalence of OF (50% to 42%) and an increase in ABD (16% to 25%), with the prevalence of osteoporosis (44% to 49%) and Al accumulation (38% to 36%) almost maintained over time.

The high prevalence of OF compared with cohorts from Europe and USA,^7^ may reflect national disparities in treatment access, especially access to parathyroidectomy, and heterogeneous standards of quality in provided care.^14-16^ The high proportion of bone Al accumulation, especially in the North and in the Northeast, suggests the need for reinforcing strategies to avoid Al exposure, as more rigorous limits for Al concentrations in water are used for dialysis (< 3 µg/L).^17^

This study has limitations to acknowledge. This is an essentially descriptive study. Our study has strengths: it demonstrated an elevated prevalence of OF, osteoporosis, vascular calcification, clinical symptoms and regional differences in the deposition of metals in bone.

## CONCLUSIONS

In this cohort, an elevated proportion of patients were diagnosed with OF and ABD, as well as osteoporosis, vascular calcification and clinical symptoms. Regional differences in the deposition of metals in bone were detected.

## Data Availability

All data produced in the present study are available upon reasonable request to the authors

## ACKNOWLEDGMENTS

The authors acknowledge the Brazilian Society of Nephrology, M.I and C.R.P. for technical support. The authors also thank the collaboration of Brazilian nephrologists and the patients included in this study.

